# Severe outcomes and length of stay among people with schizophrenia hospitalized for COVID-19: A population-based retrospective cohort study

**DOI:** 10.1101/2024.12.11.24318876

**Authors:** Jessica Gronsbell, John Wang, Hilary Thurston, Jianhui Gao, Yaqi Shi, Anthony D. Train, Debra Butt, Andrea Gershon, Braden O’Neill, Karen Tu

**Author notes:** Correspondence* Jessica Gronsbell Address: 700 University Ave, Toronto, ON, Canada, M5G 1Z5.

## Abstract

**Background and Hypothesis:** Schizophrenia is associated with substantial physical and psychiatric comorbidities that increase the risk of severe outcomes in COVID-19 infection. However, few studies have examined the differences in care and outcomes among people with schizophrenia throughout the pandemic. We hypothesized that rates of in-hospital mortality, admission to the intensive care unit (ICU), and length of stay differed among people with and without schizophrenia.

**Study Design:** We conducted a population-based retrospective cohort study using administrative health data from Ontario, Canada that included individuals hospitalized for COVID-19 between February 2020 and October 2023. We compared mortality, ICU admission, and length of stay using regression models adjusted for age, sex, comorbidities, vaccination status, and sociodemographic characteristics.

**Study Results:** We evaluated 66,959 hospital admissions, 4.3% (2,884) of which involved people with schizophrenia. People with schizophrenia had a significantly decreased rate of ICU admission (adjusted OR: 0.74, [0.67, 0.82]), a longer length of stay (adjusted RR: 1.25, [1.21, 1.30]), but a similar risk of mortality (adjusted OR: 1.09, [0.98, 1.22]) as people without schizophrenia. Age modified the relationship between schizophrenia and ICU admission. People with schizophrenia aged 60-75 were substantially less likely to be admitted to the ICU relative to those without (18.4% vs. 26.5%, p < .001).

**Conclusions:** Our findings underscore disparities in care among people with and without schizophrenia. These disparities vary by age and suggest that people with schizophrenia may not be receiving the same level of care as people without schizophrenia hospitalized for COVID-19.

## Introduction

The COVID-19 pandemic exposed significant inequities and limitations in healthcare infrastructure, particularly in crisis response for vulnerable populations [1,2]. People with schizophrenia are of particular concern due to their substantial physical and psychiatric comorbidities, which can adversely impact health, limit access to care, and reduce life expectancy [3]. Given the complexity of schizophrenia and the need for specialized care, it is critical to evaluate COVID-19 outcomes and the adequacy of care in this population [4].

A small number of studies investigated the impact of COVID-19 on people with schizophrenia early on in the pandemic [1]. Population-based studies in the U.S. and South Korea identified that people with schizophrenia were more likely to test positive for COVID-19 [2–4]. An Israeli study later found that this population is less likely to be vaccinated, placing them at higher risk for hospitalization and severe outcomes such as intensive care unit (ICU) admission and mortality [5–9]. A US study confirmed a higher COVID-19 mortality rate among people with schizophrenia, while studies from Israel, Denmark, and Spain reported increased risks of both hospitalization and COVID-19 related death in this population [4,9–12].

More recently, a study among US veterans found that people with schizophrenia are at increased risk of mortality following a COVID-19 diagnosis relative to people without schizophrenia, even after adjusting for health and treatment factors, while a study in Brazil found smaller differences in mortality compared to existing US and European evaluations [13,14]. A UK study showed that even after adjusting for vaccination status, people with schizophrenia are more likely to experience COVID-19 mortality than people without schizophrenia [15]. However, the discrepancy is significantly attenuated when adjusting for underlying health and demographic factors. A small South Korean study also did not find an increased mortality risk among people with schizophrenia [3]. Notably, only one study conducted in France examined both mortality and ICU admission to explore disparities in health outcomes and care between people with and without schizophrenia [16]. This study confirmed the higher COVID-19 mortality risk observed in other studies and highlighted a significant disparity in care as people with schizophrenia were less likely to be admitted to the ICU compared to people without schizophrenia.

These findings point to a pressing health equity crisis, as evidence indicates that people with schizophrenia may be more likely to experience severe COVID-19 infection while potentially receiving different treatment and monitoring. However, most existing studies focus on mortality rates and hospital admissions during the first year of the pandemic and therefore involve fewer than 1,000 individuals with schizophrenia who contracted COVID-19, do not account for vaccination effects, and often neglect other important measures of severity and care quality, such as length of stay and ICU admission. In many countries, most COVID-19 hospitalizations occurred after this initial period (e.g., see Supplementary Figure 1) [17]. Therefore, a study examining severe COVID-19 outcomes throughout the entire course of the pandemic is necessary to gain a more comprehensive understanding of potential health disparities impacting people with schizophrenia.

To address this gap, we conducted a study using administrative health data from Ontario, Canada, to determine whether severe outcomes and length of stay differed between individuals with and without schizophrenia between February 2020 and October 2023. Our primary objective was to compare rates of severe outcomes, including in-hospital mortality and ICU admission, while adjusting for key confounding variables such as vaccination status, comorbidities, and sociodemographic characteristics. Our secondary objective was to analyze length of hospital stay, as it serves as an indicator of infection severity and the effectiveness of hospital care.

## Methods

### Study design and data sources

We used administrative health data from the Ontario Health Data Platform (OHDP) to conduct a retrospective population-based cohort study. OHDP is a federated high-performance computing environment for secure, accurate, and privacy-protective linkage of large health datasets and was funded by Ontario’s agency for health system planning and implementation (Ontario Health) in 2021. We included all people hospitalized for COVID-19 between February 2020 and October 2023 aged 18 and older. All hospital services are covered by the Ontario Health Insurance Plan (OHIP), which is a government-run healthcare plan in the province of Ontario for all permanent residents.

Hospitalizations were identified based on admitting diagnosis codes related to COVID-19 (International Classification of Diseases (ICD)-10 codes: U07.10 or U07.12 or U07.14). A previous study of hospitalizations in Ontario found these codes to have a sensitivity of 97.8% and specificity of 99.5% [18]. We kept the first hospitalization for each patient for analysis as very few people had more than 1 COVID-19 related admission (< 4%). To identify people with schizophrenia, we used a validated case identification algorithm based on physician visits and inpatient hospitalizations that has a sensitivity of 80.1% and specificity of 68.0% [19]. People were classified as having schizophrenia if they had at least 3 ICD-9 or ICD-10 codes of schizophrenia or psychosis over a 36 month period or at least one inpatient hospitalization with that diagnosis code prior to their hospital admission.

### Outcome and covariate definitions

We considered three outcomes measuring COVID-19 severity and effectiveness of hospital care: in-hospital mortality, admission to the ICU, and length of stay. We obtained several clinical and sociodemographic covariates describing our population, including age (<60, 60-75, >75 years), sex, Charlson Comorbidity Index (CCI) score, income quintile based on residential neighborhood, and vaccination status. People were considered fully vaccinated for COVID-19 if they had received their most recent dose that qualified them as fully vaccinated (either 2 doses or 1 dose, depending on the vaccine) or a subsequent booster vaccine at least 14 days, but no more than 6 months, prior to hospital admission. In contrast, people were considered as unvaccinated if they had never received a vaccine, or if they received only the first dose of a 2-dose series vaccine less than 14 days before hospital admission, or the most recent dose they received was more than 6 months prior to hospital admission. Anyone who did not meet the criteria for fully vaccinated or unvaccinated was considered to be partially vaccinated [20].

We included these covariates in our analysis as they likely confound the relationship between schizophrenia and each of the three outcomes. Additionally, the CCI score serves as a proxy for a patient’s overall comorbidity burden which can contribute to illness severity at admission while income quintile serves as a proxy for socioeconomic status [21]. Vaccination provides protective effects against severe COVID-outcomes and has also been shown to correlate with healthcare engagement [22].

### Statistical analysis

We summarized outcomes and covariates using descriptive statistics, including means for continuous or count variables and counts and percentages for categorical variables. We fit a series of regression models to understand the relationship between schizophrenia and each of the three outcomes. The first model (Model 1) was unadjusted for any covariates, the second model (Model 2) adjusted for age, sex, income quintile, vaccination status, CCI, and the third model (Model 3) adjusted for the same covariates as Model 2 and included an interaction term between age and schizophrenia. The interaction was used to determine if the relationship between schizophrenia and the outcomes varied by age as previous work has demonstrated a strong relationship between COVID-19 severity and age [23]. Logistic regression was used to model the two binary outcomes (in-hospital mortality and ICU admission) and negative binomial regression was used to model length of stay which was an overdispersed count variable. A significance threshold of p < .05 was used. All analyses were performed in R (version 4.3.2).

### Ethics

Ethical approval was obtained from the North York General Hospital Research Ethics Board with approval #2023-0256-423.

## Results

### Population characteristics

During the study period, 66,959 people were admitted to the hospital for COVID-19 and 2,884 (4.3%) had schizophrenia (see Table 1). People with schizophrenia were more likely to be male (52.5% vs. 47.5%, p = .006), younger (70 vs. 74 years, p < .001), and be in the lowest income quintile (41.6% vs. 30.6%, p < .001). They also had higher CCI scores (41.9% vs. 32.0% for a score of 2+, p < .001), but were less likely to be unvaccinated (75.9% vs. 78.5%, p = .002).

**Table 1:** Sociodemographic, clinical, and outcome variables used in our study among people with and without schizophrenia (SCZ).

|  | Total | People with SCZ | People without SCZ |  |
| --- | --- | --- | --- | --- |
|  | *n* = 66,959 | *n* = 2,884 | *n* = 64,075 |  |
| Characteristics | *n* (%) | *n* (%) | *n* (%) | *P* value |
| Sociodemographic data |  |  |  |  |
| Age |  |  |  | **<.001** |
| <60 | 16,159(24.1%) | 736(25.5%) | 15,423(24.1%) |  |
| 60-75 | 20,302(30.3%) | 1,106(38.3%) | 19,196(30.0%) |  |
| >75 | 30,498(45.5%) | 1,042(36.1%) | 29,456(46.0%) |  |
| Sex |  |  |  | **.006** |
| Male | 36,860(55.0%) | 1,515(52.5%) | 35,345(55.2%) |  |
| Female | 30,099(45.0%) | 1,369(47.5%) | 28,730(44.8%) |  |
| Income |  |  |  | **<.001** |
| First quintile | 20,806(31.1%) | 1,201(41.6%) | 19,605(30.6%) |  |
| Second quintile | 15,098(22.5%) | 645(22.4%) | 14,453(22.6%) |  |
| Third quintile | 11,977(17.9%) | 424(14.7%) | 11,553(18.0%) |  |
| Fourth quintile | 10,267(15.3%) | 330(11.4%) | 9,937(15.5%) |  |
| Fifth quintile | 8,713(13.0%) | 280(9.7%) | 8,433(13.2%) |  |
| Clinical data at baseline |  |  |  |  |
| Charlson Comorbidity Index score |  |  |  | **<.001** |
| 0 | 32,599(48.7%) | 1,134(39.3%) | 31,465(49.1%) |  |
| 1 | 12,643(18.9%) | 542(18.8%) | 12,101(18.9%) |  |
| ≥2 | 21,717(32.4%) | 1,208(41.9%) | 20,509(32.0%) |  |
| Vaccine |  |  |  | **.002** |
| Unvaccinated | 52,494(78.4%) | 2,188(75.9%) | 50,306(78.5%) |  |
| Partially vaccinated | 1,962(2.9%) | 87(3.0%) | 1,875(2.9%) |  |
| Fully vaccinated | 12,503(18.7%) | 609(21.1%) | 11,894(18.6%) |  |
| Outcomes |  |  |  |  |
| In-hospital mortality |  |  |  |  |
| No | 57,065(85.2%) | 2,418(83.8%) | 54,647(85.3%) | **.035** |
| Yes | 9,894(14.8%) | 466(16.2%) | 9,428(14.7%) |  |
| ICU admission |  |  |  | **<.001** |
| No | 54,367(81.2%) | 2,421(83.9%) | 51,946(81.1%) |  |
| Yes | 12,592(18.8%) | 463(16.1%) | 12,129(18.9%) |  |
| Length of Stay |  |  |  | **.856** |
| 1 | 64,440(96.2%) | 2,771(96.1%) | 61,669(96.2%) |  |
| 2 | 2,401(3.6%) | 107(3.7%) | 2,294(3.6%) |  |
| ≥3 | 118(0.2%) | 6(0.2%) | 112(0.2%) |  |

### In-hospital mortality

During the study period, a total of 9,894 (14.8%) patients died in-hospital due to COVID-19. 16.2% of people with schizophrenia died, while 14.7% of people without schizophrenia died (Model 1 unadjusted Odds Ratio (OR): 1.12; 95% CI: [1.01, 1.24]; see Table 2). This relationship is potentially explained by the clinical and sociodemographic covariates that we considered, including age, sex, CCI score, income quintile, and vaccination status, as the adjusted OR for schizophrenia was further attenuated (Model 2 adjusted OR: 1.09; 95% CI: [0.98, 1.22]). The relationship between mortality and schizophrenia status was not modified by age as no significant interaction between age and schizophrenia was observed in Model 3 (Table 2).

**Table 2:** Regression modeling results for in-hospital mortality among people admitted to the hospital admitted for COVID-19 (*n* = 66,959).

| Variable | Model 1 |  | Model 2 |  | Model 3 |  |
| --- | --- | --- | --- | --- | --- | --- |
|  | OR (95% CI) | *P* value | OR (95% CI) | *P* value | OR (95% CI) | *P* value |
| Schizophrenia (SCZ) | 1.12 (1.01, 1.24) | **.033** | 1.09 (0.98, 1.22) | .095 | 1.05 (0.77, 1.45) | .742 |
| Age |  |  |  |  |  |  |
| 60 -75 years |  |  | 2.59 (2.39, 2.81) | **<.001** | 2.58 (2.37, 2.81) | **<.001** |
| > 75 years |  |  | 3.96 (3.66, 4.28) | **<.001** | 3.95 (3.65, 4.28) | **<.001** |
| Sex |  |  |  |  |  |  |
| Female |  |  | 0.76 (0.73, 0.79) | **<.001** | 0.76 (0.73, 0.79) | **<.001** |
| Income |  |  |  |  |  |  |
| Second quintile |  |  | 0.99 (0.94, 1.05) | .827 | 0.99 (0.94, 1.05) | .826 |
| Third quintile |  |  | 0.92 (0.87, 0.99) | **.019** | 0.92 (0.87, 0.99) | **.019** |
| Fourth quintile |  |  | 0.89 (0.83, 0.96) | **.001** | 0.89 (0.83, 0.96) | **.001** |
| Fifth quintile |  |  | 0.82 (0.76, 0.89)) | **<.001** | 0.82 (0.76, 0.89) | **<.001** |
| CCI score |  |  |  |  |  |  |
| 1 |  |  | 1.71 (1.60, 1.82) | **<.001** | 1.71 (1.60, 1.82) | **<.001** |
| $\geq 2$ | | | 2.35 (2.23, 2.48) | **<.001** | 2.35 (2.23, 2.48) | **<.001** |
| Vaccine |  |  |  |  |  |  |
| Partially vaccinated |  |  | 1.12 (0.99, 1.27) | .060 | 1.12 (1.00, 1.27) | .060 |
| Fully vaccinated |  |  | 0.54 (0.51, 0.57) | **<.001** | 0.54 (0.51, 0.57) | **<.001** |
| Age × SCZ | | | | | | |
| 60 - 75 years |  |  |  |  | 1.12 (0.70, 1.82) | .060 |
| > 75 years |  |  |  |  | 1.08 (0.67, 1.73) | .751 |

Among the covariates, vaccination status was one of the strongest factors influencing mortality, with an adjusted OR for being fully vaccinated relative to being unvaccinated of 0.54 (95% CI: [0.51,0.57]; Model 2). Several other covariates were strongly associated with mortality, including older age (Model 2 adjusted OR: 2.59; 95% CI: [2.39, 2.81] for age 60-75 relative to age < 60; adjusted OR: 3.96; 95% CI: [3.66, 4.28] for age > 75 relative to age < 60), a CCI score above 2 (Model 2 adjusted OR: 2.35; 95% CI: [2.23, 2.48)], a higher income quintile (Model 2 adjusted OR: 0.82; 95% CI: [0.76, 0.89]), and female sex (Model 2 adjusted OR: 0.76; 95% CI: [0.73, 0.79]). The findings are similar for the model including the interaction term for age and schizophrenia status (Model 3).

### Intensive care unit admission

A total of 12,592 (18.8%) people were admitted to the ICU. People with schizophrenia were less likely to be admitted to the ICU:16.1% of people with schizophrenia were admitted to the ICU compared to 18.9% of people without schizophrenia (Model 1 unadjusted OR: 0.82; 95% CI: [0.74, 0.91]; see Table 3). This association remained significant in Model 2, which adjusted for confounding variables, suggesting that the observed difference in ICU admission cannot be explained by the sociodemographic and clinical factors considered in our study (Model 2 adjusted OR: 0.74; 95% CI: [0.67, 0.82]). Additionally, there was a significant interaction between age and schizophrenia, indicating the difference in ICU admission is more pronounced for people above 60 years of age. Relative to people under age 60 (Model 3 adjusted OR: 0.95; 95% CI: [0.79, 1.13]), the discrepancy in ICU admission for people between the ages of 60 and 75 is significant (Model 3 adjusted OR: 0.63; 95% CI: [0.47, 0.84]) and marginally significant for people over age 75 (Model 3 adjusted OR: 0.71; 95% CI: [0.51, 0.99]).

**Table 3:** Regression modeling results for ICU admission among people admitted to the hospital admitted for COVID-19 (*n* = 66,959).

| Variable | Model 1 |  | Model 2 |  | Model 3 |  |
| --- | --- | --- | --- | --- | --- | --- |
|  | OR (95% CI) | *P* value | OR (95% CI) | *P* value | OR (95% CI) | *P* value |
| Schizophrenia (SCZ) | 0.82 (0.74, 0.91) | **<.001** | 0.74 (0.67, 0.82) | .095 | 0.95 (0.79, 1.13) | .536 |
| Age |  |  |  |  |  |  |
| 60 -75 years |  |  | 1.06 (1.01, 1.12) | **.014** | 1.08 (1.03, 1.14) | **.002** |
| > 75 years |  |  | 0.38 (0.36, 0.40) | **<.001** | 0.39 (0.36, 0.41) | **<.001** |
| Sex |  |  |  |  |  |  |
| Female |  |  | 0.74 (0.71, 0.77) | **<.001** | 0.74 (0.71, 0.77) | **<.001** |
| Income |  |  |  |  |  |  |
| Second quintile |  |  | 0.98 (0.93, 1.04) | .478 | 0.98 (0.93, 1.04) | .501 |
| Third quintile |  |  | 0.95 (0.90, 1.01) | .097 | 0.95 (0.90, 1.01) | .107 |
| Fourth quintile |  |  | 0.90 (0.84, 0.95) | **.001** | 0.90 (0.84, 0.95) | **.001** |
| Fifth quintile |  |  | 0.95 (0.89, 1.01) | .101 | 0.95 (0.89, 1.01) | .108 |
| CCI score |  |  |  |  |  |  |
| 1 |  |  | 1.44 (1.37, 1.52) | **<.001** | 1.44 (1.37, 1.52) | **<.001** |
| $\geq 2$ | | | 1.19 (1.13, 1.25) | **<.001** | 1.19 (1.13, 1.25) | **<.001** |
| Vaccine |  |  |  |  |  |  |
| Partially vaccinated |  |  | 1.01 (0.90, 1.14) | .820 | 1.01 (0.90, 1.13) | .875 |
| Fully vaccinated |  |  | 0.61 (0.57, 0.64) | **<.001** | 0.61 (0.57, 0.64) | **<.001** |
| Age × SCZ | | | | | | |
| 60 - 75 years |  |  |  |  | 0.63 (0.47, 0.84) | **.002** |
| > 75 years |  |  |  |  | 0.71 (0.51, 0.99) | **.046** |

With respect to the clinical and sociodemographic covariates, the adjusted OR for being fully vaccinated relative to unvaccinated is 0.61 (95% CI: [0.57, 0.64]; Model 2). Vaccination status is one of the strongest covariates influencing ICU admission and is accompanied by age (Model 2 adjusted OR: 0.38; 95% CI: [0.36, 0.40] for age 60-75 relative to age < 60; adjusted OR: 1.06; 95% CI: [1.01, 1.12] for age > 75 relative to age < 60), a CCI score above 1 (Model 2 adjusted OR: 1.44; 95% CI: [1.37, 1.52]; Model 2), and female sex (Model 2 adjusted OR: 0.74; 95% CI: [0.71, 0.77]). Higher income is associated with a slightly lower risk of ICU admission, but not consistently across the income quintiles. These patterns are similar for the model that includes the interaction term between age and schizophrenia status (Model 3).

### Length of stay

The overall average length of stay was 12.00 days. People with schizophrenia stayed an average of 15.39 days in hospital, while people without schizophrenia stayed an average of 11.85 days (Model 1 unadjusted RR: 1.30; 95% CI: [1.25, 1.35], see Table 4). The relationship between length of stay and schizophrenia was significant after adjusting for all of the covariates (Model 2 adjusted RR: 1.25, 95% CI: [1.21, 1.30]). Similar to the mortality outcome, age did not modify the observed association between these two variables (Model 3).

**Table 4:** Regression modeling results for length of stay among people admitted to the hospital admitted for COVID-19 (*n* = 66,959).

| Variable | Model 1 |  | Model 2 |  | Model 3 |  |
| --- | --- | --- | --- | --- | --- | --- |
|  | RR (95% CI) | *P* value | RR (95% CI) | *P* value | RR (95% CI) | *P* value |
| Schizophrenia (SCZ) | 1.30 (1.25, 1.35) | **<.001** | 1.25 (1.21, 1.30) | **<.001** | 1.34 (1.25, 1.44) | **<.001** |
| Age |  |  |  |  |  |  |
| 60 -75 years |  |  | 1.20 (1.18, 1.23) | **<.001** | 1.21 (1.18, 1.23) | **<.001** |
| > 75 years |  |  | 1.15 (1.12, 1.17) | **<.001** | 1.15 (1.13, 1.17) | **<.001** |
| Sex |  |  |  |  |  |  |
| Female |  |  | 0.92 (0.91, 0.94) | **<.001** | 0.92 (0.91, 0.94) | **<.001** |
| Income |  |  |  |  |  |  |
| Second quintile |  |  | 1.02 (1.00, 1.04) | .084 | 1.02 (1.00, 1.04) | .080 |
| Third quintile |  |  | 0.96 (0.94, 0.98) | **<.001** | 0.96 (0.94, 0.98) | **<.001** |
| Fourth quintile |  |  | 0.94 (0.92, 0.96) | **<.001** | 0.94 (0.92, 0.96) | **<.001** |
| Fifth quintile |  |  | 0.95 (0.93, 0.97) | **<.001** | 0.95 (0.93, 0.98) | **<.001** |
| CCI score |  |  |  |  |  |  |
| 1 |  |  | 1.25 (1.22, 1.27) | **<.001** | 1.25 (1.22, 1.27) | **<.001** |
| $\geq 2$ | | | 1.45 (1.43, 1.48) | **<.001** | 1.45 (1.43, 1.48) | **<.001** |
| Vaccine |  |  |  |  |  |  |
| Partially vaccinated |  |  | 0.96 (0.92, 1.01) | .099 | 0.96 (0.92, 1.01) | 0.093 |
| Fully vaccinated |  |  | 0.78 (0.76, 0.79) | **<.001** | 0.78 (0.76, 0.79) | **<.001** |
| Age × SCZ | | | | | | |
| 60 - 75 years |  |  |  |  | 1.25 (1.11, 1.40) | **<.001** |
| > 75 years |  |  |  |  | 1.21 (1.07, 1.35) | **.002** |

Vaccination status is a strong factor influencing length of stay, with an adjusted RR for being fully vaccinated relative to unvaccinated of 0.78 (95% CI: [0.76, 0.79]; Model 2). A CCI score above 2 (Model 2 adjusted RR: 1.45; 95% CI: [1.43, 1.48]) and older age are also significantly associated with length of stay (Model 2 adjusted RR 1.20; 95% CI: [1.18, 1.23] for age 60-75 relative to age < 60 and adjusted RR: 1.15; 95% CI: [1.12, 1.17] for age > 75 relative to age < 60). Female sex (Model 2 adjusted RR: 0.92; 95% CI: [0.91, 0.94]) and lower income quintile are associated with length of stay. These associations are similar in the model including an interaction term between age and schizophrenia status (Model 3).

## Discussion

We conducted a population-based retrospective cohort study using administrative data from Ontario, Canada, and observed health and healthcare disparities between individuals with and without schizophrenia who had COVID-19. Unlike previous research, our study spanned the first several years of the pandemic, included a much larger cohort of individuals with schizophrenia who were hospitalized for COVID-19, accounted for vaccination effects, and considered outcomes related to both COVID-19 severity and care. The results of our regression analyses adjusted for key clinical and sociodemographic variables showed people with schizophrenia (i) are no more likely to experience in-hospital mortality, (ii) have significantly longer hospital stays, but (iii) are much less frequently admitted to the ICU. Furthermore, the gap in ICU admission is particularly pronounced among people over age 60. Our findings regarding ICU admission align with a French study and suggest that people with schizophrenia may not be receiving the same care as people without schizophrenia [16].

More specifically, the French study was conducted between February and June 2020 and found that people with schizophrenia are more likely to experience in-hospital mortality, but are much less frequently admitted to the ICU. As this period was before widespread vaccination efforts, this study could not adjust for the vaccine effects. A study conducted in Ontario in 2021 found that people with schizophrenia were more likely to be unvaccinated relative to people without schizophrenia and that this discrepancy was partially attributable to lower healthcare service use [22]. We similarly found that people with schizophrenia were more likely to be unvaccinated within the OHDP database through June 2023; 20.8% of individuals with schizophrenia were unvaccinated compared to 11.5% of those without schizophrenia. However, among our study population which only included people hospitalized for COVID-19, people with schizophrenia were actually less likely to be unvaccinated relative to people without schizophrenia (75.9% vs. 78.5%, p = .002).

The higher vaccination rate among people with schizophrenia may be one of the many contributing factors to the attenuated risk of mortality in our analyses relative to the French study, as well as previous literature that evaluates vaccination rates across entire regions or among people with a COVID-19 diagnosis, but not necessarily a hospitalization. The higher vaccination rate among those with schizophrenia hospitalized for COVID-19 is also consistent with prior work. For example, a recent meta-analysis found that while vaccination does substantially decrease the risk of mortality among people with severe mental illness, this population is still two times more likely to be hospitalized relative to people without severe mental illness even after adjusting for vaccination [24].

We also found that the association between schizophrenia and in-hospital mortality is further attenuated when adjusting for the covariates considered. This may be due to the underlying differences among people with and without schizophrenia across sex, age, comorbidity burden, vaccination status, and income, which are all factors associated with schizophrenia and mortality from COVID-19 [25–27]. However, we did not observe effect modification by age like the French study. One potential explanation is that the mean age in our study population is 73 years of age, which is older than the previous study. It may be possible that increased mortality exists among younger populations, but our sample size did not permit a reliable stratified analysis to confirm this hypothesis.

In contrast to in-hospital mortality, the relationships between schizophrenia and ICU admission, as well as length of stay, are not explained by the covariates considered. Our adjusted analyses point to a similar effect size for ICU admission as the French study and an increased length of stay among those with schizophrenia. With respect to ICU admission, the French study found the most notable differences among people over age 55. Our findings are similar in that individuals over 60 years of age with schizophrenia are much less frequently admitted to the ICU relative to individuals without schizophrenia. One potential explanation of this finding is that older patients may be more frequently referred from hospitals or institutions than patients without a diagnosis of severe mental illness, though our data does not contain information to confirm this hypothesis. It is likely that institutionalization is a risk factor for COVID-19 severe infection in elderly patients and may partially explain this finding [5,16].

Another difference between the present study and the French study, is that the authors had access to a small number of in-hospital variables that measured disease severity, including the SAPS II score at ICU admission, recourse to invasive mechanical ventilation, and recourse to continuous renal-replacement therapy. However, even after adjusting for these factors, the relationship between schizophrenia and ICU admission persisted. The authors speculated that additional factors beyond the covariates considered, many of which are traditionally not available within administrative health data, could explain this finding. We similarly hypothesize that our findings can be contextualized within extensive literature exploring the factors related to disparities in health and care outcomes between people with and without schizophrenia [28,29].

In particular, people with schizophrenia are considered a vulnerable population for many reasons, including risk taking behaviors, living conditions, barriers to healthcare access and preventative measures, and psychiatric and physical comorbidities. These factors can significantly affect health outcomes and healthcare service use. A recent study reported that individuals with schizophrenia have a higher likelihood of contracting COVID-19 due to reduced adherence to mask-wearing and stay-at-home orders, lower risk awareness, and living in crowded congregate settings [30]. Another study suggested that increased COVID-19 mortality among those with schizophrenia might be attributed to congregate living conditions rather than the illness itself [31]. Additionally, individuals with schizophrenia use outpatient services less frequently than those without schizophrenia, potentially leading to delays in COVID-19 diagnosis and treatment [32]. Reduced access to pre-admission medical care may have been further exacerbated with the shift from in-person to virtual care during the pandemic. Our recent study using University of Toronto Practice-Based Research Network (UTOPIAN) primary care electronic medical record data found that preventive care was substantially disrupted for people with schizophrenia during the shift to virtual care [33]. Furthermore, among the many comorbidities affecting people with schizophrenia, cardiovascular disease is the leading cause of death [34]. It is well-established that individuals with cardiovascular disease face a higher risk of severe COVID-19 infection and mortality [35]. Existing evidence also points to higher rates of respiratory illnesses among individuals with schizophrenia, such as emphysema and COPD, as well as higher rates of obesity [4,36–41]. Similar to cardiovascular disease, these conditions confer a higher risk of severe COVID-19 infection and mortality. Psychiatric comorbidities can further increase the risk of severe COVID-19 outcomes and contribute to increased hospital admission rates among people with schizophrenia [2]. Smoking is also more prevalent among individuals with psychiatric disorders, which is a behavior that can increase the risk of severe outcomes related to COVID-19 [8,40].

Several other factors may also help to explain the differences in care observed in our study, particularly the differences in ICU admission and length of stay between people with and without schizophrenia in spite of their similar in-hospital mortality rates. One potential explanation is reduced access to pre-admission medical care among people with schizophrenia, which could contribute to increased hospitalization [31]. On the other hand, people with schizophrenia, who in our study population were more likely to be in a lower income quintile and who often have lower executive function, may be less able to provide self-care at home. They may therefore be admitted to the hospital with less severe COVID-19 relative to people without schizophrenia [4,31]. Lower disease severity, together with higher vaccination rates, is one possible explanation to the lower ICU admission rates observed in our study. Additionally, people with schizophrenia may receive lower quality of care due to both patient (e.g., fear, cognitive limitations, socioeconomic disadvantage) and provider (e.g., discomfort, lack of training) factors, which may be another contributing factor to the disparities in ICU admission and length of stay [42].

As noted by Fond et al., our findings regarding ICU admission may also be an example of the conflict between utility-based and equity-based arguments in healthcare [16]. People with schizophrenia have some of the poorest prognosis factors that justify ICU admission, including functional status, do-not-resuscitate orders, and their referring facility [43]. Many of these factors are not modifiable and may contribute to our finding that people with schizophrenia are less frequently admitted to ICU in spite of having longer lengths of stay [44]. It is well known that coordination challenges between hospital and psychiatric care teams can lead to delays in access to care, and that stigma and communication barriers further impact individuals with schizophrenia [45,46]. Together with additional psychiatric management, it is also possible that these challenges lead to their longer lengths of stay. Further quantitative and qualitative research is needed to disentangle the clinical and social factors that contribute to the observed disparities in outcomes and care between individuals with and without schizophrenia.

### Limits and Perspectives

This study relied on data from the Ontario Health Data Platform, which is an administrative health database developed by the Ontario government in response to the COVID-19 pandemic. The data begins on January 1, 2010 and we therefore may have missed schizophrenia diagnoses that occurred before this date. While this could reduce the observed disparities, it is likely that individuals with schizophrenia who are missed by our case identification algorithm are sufficiently stable to avoid interaction with the healthcare system for mental health services and may not be at increased risk for severe infection. Our case identification algorithm also misclassified individuals with and without schizophrenia due to its inherent limitations, which may attenuate the observed associations in our analyses [47].

Lastly, since this study relies on observational health data, it cannot provide a complete picture of individuals’ health status or account for all potential confounding variables. For example, we did not have information on the time between onset of COVID-19 infection and hospitalization. We also did not have complete information on psychiatric management such as psychotropic medications that may be contraindicated for some COVID-19 treatments, body mass index, executive function, comorbid psychiatric disorders, pre-admission or in-hospital care, or data on immune response. All of these factors may confound the observed associations. Accounting for major COVID-19 waves or vaccination rollouts could also provide additional insights into how our findings evolved over time with changing treatments and healthcare responses. It is also unclear if our findings generalize to other settings and future research in this direction is warranted.

## Conclusion

This study points to disparities in severe outcomes and healthcare between people hospitalized for COVID-19 with and without schizophrenia. Within our study population, people with schizophrenia are no more likely to experience in-hospital mortality due to COVID-19, but are more likely to have longer stays in hospital and less likely to be admitted to the ICU, particularly among older age groups. Our findings emphasize the importance of personalized clinical management and strategies for people with schizophrenia who are infected with COVID-19.

## Data Availability

Data used in this study was from the Ontario Health Data Platform (OHDP), which consisted of record-level, coded and linkable health data sets from the population of Ontario, Canada. These data were held securely, privacy-protected for the benefit of individuals, and are not publicly available.

## Acknowledgements

The authors thank project team members M. Bianca Seaton, Lindsay Lustig, Krystle Amog, Angela Coderre-Ball, Maria Carla Lapadula, Ye Lennon Li, Dorsa Mohammadrezaei, Angela Ortigoza, Javier Silva Valencia, and Krishna Karthik Vemuri, who provided support with the following range of tasks: project management and coordination, data cleaning, methodological strategy, literature searches, and manuscript formatting.

## Funding Statement

This study received funding from the Ontario Ministry of Health (MOH). Parts of this material are based on data and information compiled and provided by the Ontario Ministry of Health (Ontario Health Data Platform). However, the analyses, conclusions, opinions and statements expressed herein are those of the author, and not necessarily those of the Ministry.

The study also received funding from the Rathlyn Foundation Primary Care EMR Research and Discovery Fund.

This research was funded in whole or in part by the Province of Ontario. Parts of this material are based on data and information compiled and provided by the Province. However, the analyses, conclusions, opinions and statements expressed herein are those of the author, and not necessarily those of the Province.

## Disclaimer

DB, KT, AG, BO, JG and AT received financial support from the First Mover’s Fund, Ontario Ministry of Health. DB, KT, and BO receive Research Investigator Awards from the Department of Family and Community Medicine at the University of Toronto. KT receives a Chair in Family and Community Medicine Research in Primary Care at University Health Network.

## Supplementary Materials

**Supplementary Figure 1:**
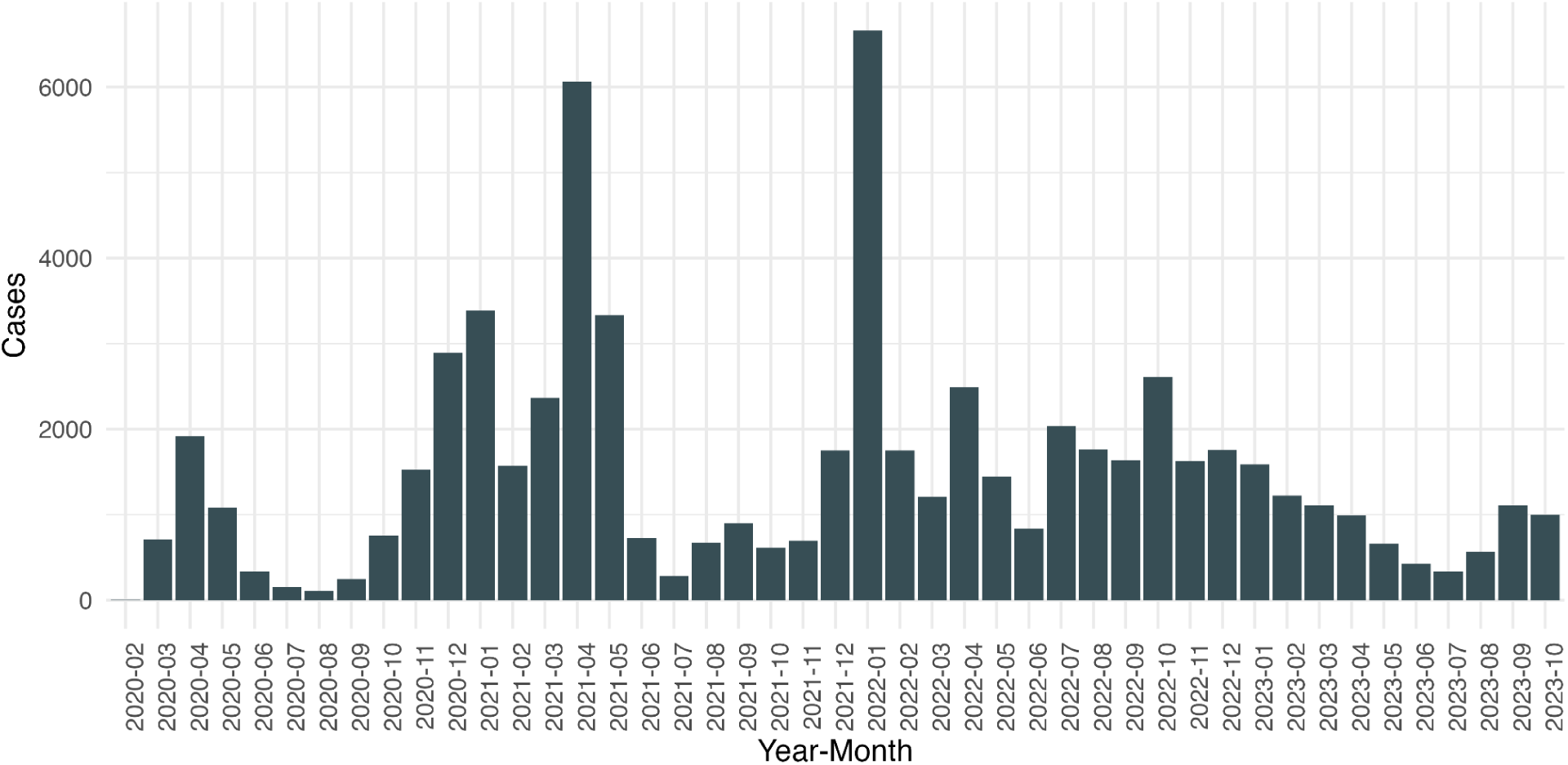
Monthly count of COVID-19 hospitalizations between February, 2020 and October, 2023 in Ontario, Canada.

## Notes

### Author Declarations

Ethics committee/IRB of the North York General Hospital Research Ethics Board gave ethical approval for this work.

### Summary of Updates

update some results according to feedbacks from reviewers

## References

1. Pardamean E, Roan W, Iskandar KTA, Prayangga R, Hariyanto TI. Mortality from coronavirus disease 2019 (Covid-19) in patients with schizophrenia: A systematic review, meta-analysis and meta-regression. Gen Hosp Psychiatry. 2022;75: 61–67.

2. Wang Q, Xu R, Volkow ND. Increased risk of COVID-19 infection and mortality in people with mental disorders: analysis from electronic health records in the United States. World Psychiatry. 2021;20: 124–130.

3. Ji W, Huh K, Kang M, Hong J, Bae GH, Lee R, et al. Effect of Underlying Comorbidities on the Infection and Severity of COVID-19 in Korea: a Nationwide Case-Control Study. J Korean Med Sci. 2020;35: e237.

4. Tzur Bitan D, Krieger I, Kridin K, Komantscher D, Scheinman Y, Weinstein O, et al. COVID-19 Prevalence and Mortality Among Schizophrenia Patients: A Large-Scale Retrospective Cohort Study. Schizophr Bull. 2021;47: 1211–1217.

5. Fond G, Nemani K, Etchecopar-Etchart D, Loundou A, Goff DC, Lee SW, et al. Association Between Mental Health Disorders and Mortality Among Patients With COVID-19 in 7 Countries: A Systematic Review and Meta-analysis. JAMA Psychiatry. 2021;78: 1208–1217.

6. Toubasi AA, AbuAnzeh RB, Tawileh HBA, Aldebei RH, Alryalat SAS. A meta-analysis: The mortality and severity of COVID-19 among patients with mental disorders. Psychiatry Res. 2021;299: 113856.

7. Vai B, Mazza MG, Delli Colli C, Foiselle M, Allen B, Benedetti F, et al. Mental disorders and risk of COVID-19-related mortality, hospitalisation, and intensive care unit admission: a systematic review and meta-analysis. Lancet Psychiatry. 2021;8: 797–812.

8. Karaoulanis SE, Christodoulou NG. Do patients with schizophrenia have higher infection and mortality rates due to COVID-19? A systematic review. Psychiatriki. 2021;32: 219–223.

9. Goldberger N, Bergman-Levy T, Haklai Z, Yoffe R, Davidson M, Susser E, et al. COVID-19 and severe mental illness in Israel: testing, infection, hospitalization, mortality and vaccination rates in a countrywide study. Mol Psychiatry. 2022;27: 3107–3114.

10. Moreno-Juste A, Poblador-Plou B, Ortega-Larrodé C, Laguna-Berna C, González-Rubio F, Aza-Pascual-Salcedo M, et al. Mental health and risk of death and hospitalization in COVID-19 patients. Results from a large-scale population-based study in Spain. PLoS One. 2024;19: e0298195.

11. Nemani K, Li C, Olfson M, Blessing EM, Razavian N, Chen J, et al. Association of Psychiatric Disorders With Mortality Among Patients With COVID-19. JAMA Psychiatry. 2021;78: 380–386.

12. Reilev M, Kristensen KB, Pottegård A, Lund LC, Hallas J, Ernst MT, et al. Characteristics and predictors of hospitalization and death in the first 11 122 cases with a positive RT-PCR test for SARS-CoV-2 in Denmark: a nationwide cohort. Int J Epidemiol. 2020;49: 1468–1481.

13. Bowersox NW, Browne J, Grau PP, Merrill SL, Haderlein TP, Llorente MD, et al. COVID-19 mortality among veterans with serious mental illness in the veterans health administration. J Psychiatr Res. 2023;163: 222–229.

14. Costa DFB, Rossignoli P, Pontarolli DRS, Junior PC, Assolari CL, Nasr AMLF, et al. Increased COVID-19 mortality in patients with schizophrenia: A retrospective study in Brazil. Schizophr Res. 2024;271: 200–205.

15. Hassan L, Sawyer C, Peek N, Lovell K, Carvalho AF, Solmi M, et al. Heightened COVID-19 Mortality in People With Severe Mental Illness Persists After Vaccination: A Cohort Study of Greater Manchester Residents. Schizophr Bull. 2023;49: 275–284.

16. Fond G, Pauly V, Leone M, Llorca P-M, Orleans V, Loundou A, et al. Disparities in Intensive Care Unit Admission and Mortality Among Patients With Schizophrenia and COVID-19: A National Cohort Study. Schizophr Bull. 2021;47: 624–634.

17. Dong E, Ratcliff J, Goyea TD, Katz A, Lau R, Ng TK, et al. The Johns Hopkins University Center for Systems Science and Engineering COVID-19 Dashboard: data collection process, challenges faced, and lessons learned. Lancet Infect Dis. 2022;22: e370–e376.

18. Moura CS, Neville A, Liao F, Wen B, Razak F, Roberts S, et al. Validity of hospital diagnostic codes to identify SARS-CoV-2 infections in reference to polymerase chain reaction results: a descriptive study. CMAJ Open. 2023;11: E982–E987.

19. Kurdyak P, Lin E, Green D, Vigod S. Validation of a Population-Based Algorithm to Detect Chronic Psychotic Illness. Can J Psychiatry. 2015;60: 362–368.

20. Tenforde MW, Self WH, Adams K, Gaglani M, Ginde AA, McNeal T, et al. Association Between mRNA Vaccination and COVID-19 Hospitalization and Disease Severity. JAMA. 2021;326: 2043–2054.

21. Christensen DM, Strange JE, Gislason G, Torp-Pedersen C, Gerds T, Fosbøl E, et al. Charlson Comorbidity Index Score and Risk of Severe Outcome and Death in Danish COVID-19 Patients. J Gen Intern Med. 2020;35: 2801–2803.

22. Kurdyak P, Lebenbaum M, Patrikar A, Rivera L, Lu H, Scales DC, et al. SARS-CoV-2 vaccination prevalence by mental health diagnosis: a population-based cross-sectional study in Ontario, Canada. CMAJ Open. 2023;11: E1066–E1074.

23. Romero Starke K, Reissig D, Petereit-Haack G, Schmauder S, Nienhaus A, Seidler A. The isolated effect of age on the risk of COVID-19 severe outcomes: a systematic review with meta-analysis. BMJ Glob Health. 2021;6. doi:10.1136/bmjgh-2021-006434

24. Dang W, Long I, Zhao Y, Xiang Y-T, Smith RD. Effectiveness of COVID-19 Vaccines in People with Severe Mental Illness: A Systematic Review and Meta-Analysis. Vaccines (Basel). 2024;12. doi:10.3390/vaccines12091064

25. Peres IT, Bastos LSL, Gelli JGM, Marchesi JF, Dantas LF, Antunes BBP, et al. Sociodemographic factors associated with COVID-19 in-hospital mortality in Brazil. Public Health. 2021;192: 15–20.

26. Drefahl S, Wallace M, Mussino E, Aradhya S, Kolk M, Brandén M, et al. A population-based cohort study of socio-demographic risk factors for COVID-19 deaths in Sweden. Nat Commun. 2020;11: 5097.

27. de Lusignan S, Dorward J, Correa A, Jones N, Akinyemi O, Amirthalingam G, et al. Risk factors for SARS-CoV-2 among patients in the Oxford Royal College of General Practitioners Research and Surveillance Centre primary care network: a cross-sectional study. Lancet Infect Dis. 2020;20: 1034–1042.

28. Walker ER, McGee RE, Druss BG. Mortality in mental disorders and global disease burden implications: a systematic review and meta-analysis. JAMA Psychiatry. 2015;72: 334–341.

29. Eack SM, Newhill CE, Anderson CM, Rotondi AJ. Quality of life for persons living with schizophrenia: more than just symptoms. Psychiatr Rehabil J. 2007;30: 219–222.

30. Pinkham AE, Ackerman RA, Depp CA, Harvey PD, Moore RC. COVID-19-related psychological distress and engagement in preventative behaviors among individuals with severe mental illnesses. NPJ Schizophr. 2021;7: 7.

31. Kozloff N, Mulsant BH, Stergiopoulos V, Voineskos AN. The COVID-19 Global Pandemic: Implications for People With Schizophrenia and Related Disorders. Schizophr Bull. 2020;46: 752–757.

32. Zhu JM, Myers R, McConnell KJ, Levander X, Lin SC. Trends In Outpatient Mental Health Services Use Before And During The COVID-19 Pandemic. Health Aff . 2022;41: 573–580.

33. Stephenson E, Yusuf A, Gronsbell J, Tu K, Melamed O, Mitiku T, et al. Disruptions in Primary Care among People with Schizophrenia in Ontario, Canada, During the COVID-19 Pandemic. Can J Psychiatry. 2022; 7067437221140384.

34. Olfson M, Gerhard T, Huang C, Crystal S, Stroup TS. Premature Mortality Among Adults With Schizophrenia in the United States. JAMA Psychiatry. 2015;72: 1172–1181.

35. Aggarwal G, Cheruiyot I, Aggarwal S, Wong J, Lippi G, Lavie CJ, et al. Association of Cardiovascular Disease With Coronavirus Disease 2019 (COVID-19) Severity: A Meta-Analysis. Curr Probl Cardiol. 2020;45: 100617.

36. Sánchez-Rico M, Limosin F, Hoertel N. Is a Diagnosis of Schizophrenia Spectrum Disorder Associated With Increased Mortality in Patients With COVID-19? Am J Psychiatry. 2022;179: 71–73.

37. Mohan M, Perry BI, Saravanan P, Singh SP. COVID-19 in People With Schizophrenia: Potential Mechanisms Linking Schizophrenia to Poor Prognosis. Front Psychiatry. 2021;12: 666067.

38. Fancourt D, Finn S. What Is the Evidence on the Role of the Arts in Improving Health and Well-Being. 2019.

39. Zareifopoulos N, Bellou A, Spiropoulou A, Spiropoulos K. Prevalence of Comorbid Chronic Obstructive Pulmonary Disease in Individuals Suffering from Schizophrenia and Bipolar Disorder: A Systematic Review. COPD. 2018;15: 612–620.

40. Fonseca L, Diniz E, Mendonça G, Malinowski F, Mari J, Gadelha A. Schizophrenia and COVID-19: risks and recommendations. Braz J Psychiatry. 2020;42: 236–238.

41. Fond G, Pauly V, Orleans V, Antonini F, Fabre C, Sanz M, et al. Increased in-hospital mortality from COVID-19 in patients with schizophrenia. Encephale. 2021;47: 89–95.

42. Institute of Medicine, Board on Population Health and Public Health Practice, Committee on HIV Screening and Access to Care. HIV Screening and Access to Care: Health Care System Capacity for Increased HIV Testing and Provision of Care. National Academies Press; 2011.

43. Ruijs CDM, Kerkhof AJFM, van der Wal G, Onwuteaka-Philipsen BD. Depression and explicit requests for euthanasia in end-of-life cancer patients in primary care in the Netherlands: a longitudinal, prospective study. Fam Pract. 2011;28: 393–399.

44. Zelle H, Kemp K, Bonnie RJ. Advance directives in mental health care: evidence, challenges and promise. World Psychiatry. 2015;14: 278–280.

45. Murch R. Improving attitudes to mental health patients in ICU. Nurs N Z. 2016;22: 30–31.

46. Weare R, Green C, Olasoji M, Plummer V. ICU nurses feel unprepared to care for patients with mental illness: A survey of nurses’ attitudes, knowledge, and skills. Intensive Crit Care Nurs. 2019;53: 37–42.

47. Yi GY. Statistical Analysis with Measurement Error or Misclassification: Strategy, Method and Application. Springer; 2017.

